# Continuous Low-Dose versus Standard-Dose Capecitabine Monotherapy as Second/Third-Line Chemotherapy for Metastatic Gastrointestinal Malignancies: A Retrospective Multicenter Analysis

**DOI:** 10.64898/2025.12.14.25342220

**Authors:** Swaroop Revannasiddaiah, Irappa Madabhavi, Kailash Chandra Pandey, Nirdosh Kumar Pant, Siddharth Vats, Priyanka Thakur, Mukesh Sharma, Anuradha Pinninti, Sridhar Papaiah Susheela, Madhup Rastogi

## Abstract

**Background:** Capecitabine monotherapy is frequently employed as second/third-line treatment for metastatic gastrointestinal malignancies. Standard dosing often produces severe toxicities incompatible with palliative care principles. This study compared continuous low-dose (CLD) versus standard-dose (StD) capecitabine in the palliative setting.

**Methods:** After at least one prior line of chemotherapy, 33 and 45 patients received StD (1.0-1.25 g/m^2^ bid on days 1-14 of a 21-day cycle) and CLD (500 mg bid daily without interruption) capecitabine, respectively, between March 2013 and August 2016 for metastatic gall bladder (n=51), pancreatic (n=14), and gastric (n=13) cancers. Toxicity and treatment discontinuation rates were compared using Fisher’s Exact Test. Progression-free survival (PFS) and overall survival (OS) were compared using the Mann-Whitney U test.

**Results:** Grade ≥3 toxicity was significantly higher in StD versus CLD (57.6% vs 4.4%; p<0.001). Treatment discontinuation due to toxicity was higher in StD versus CLD (66.7% vs 2.2%; p<0.001). Grade 5 toxicity occurred in 3 patients (9.1%) in the StD arm versus 0 in the CLD arm. Median PFS was 90 days (StD) versus 119 days (CLD; p=0.018). Median OS was 160 days (StD) versus 181 days (CLD; p=0.085). Disease stabilization was more common in CLD (51.1% vs 54.5%), while progression-free survival was prolonged.

**Conclusions:** While lower toxicities and discontinuation rates were expected in the CLD arm, the superior PFS and comparable OS with substantially reduced toxicity were striking. The metronomic low-dose approach provides a more favorable balance between disease control and tolerability in palliative gastrointestinal cancer therapy. Further prospective randomized trials are warranted.

## 1. Introduction

Capecitabine, an oral 5-fluorouracil prodrug, has become an important therapeutic option in the management of metastatic gastrointestinal malignancies, including advanced gastric, pancreatic, and gallbladder cancers.[1][2] As a second or third-line chemotherapy agent, it is commonly employed in the palliative setting where conventional cisplatin-based regimens are either contraindicated or have failed.[3] However, the administration of capecitabine at standard doses (1.0-1.25 g/m^2^ twice daily for 14 days in a 21-day cycle) frequently results in substantial toxicities, including hand-foot syndrome, severe diarrhea, and hematologic complications, which often necessitate dose reductions or treatment discontinuation.[2][4]

This toxicity burden represents a significant challenge in palliative oncology, where the primary goal is to optimize quality of life while maintaining disease control. Aggressive chemotherapy-related adverse effects can paradoxically worsen patient outcomes and contradict fundamental palliative care principles.[5] Recognition of this clinical dilemma has renewed interest in metronomic chemotherapy - the continuous administration of low-dose agents without treatment-free intervals.[6][7] Metronomic dosing offers theoretical advantages including sustained anti-angiogenic effects, reduced toxicity, and potential benefits against chemotherapy-resistant cancer stem cells, while maintaining drug exposure within the tumor microenvironment.[6]

Limited clinical evidence exists comparing the efficacy and tolerability of continuous low-dose versus standard-dose capecitabine monotherapy in the palliative setting. Given the increasing recognition that maximum tolerated doses may not be optimal in metastatic disease, and given the practical advantage of oral chemotherapy in resource-limited settings, we conducted a retrospective analysis comparing continuous low-dose capecitabine (0.5 g twice daily without interruption) to standard-dose capecitabine in patients with metastatic gastrointestinal malignancies who had received at least one prior line of chemotherapy. This pragmatic comparison aims to inform treatment selection for clinicians managing patients where both regimens represent reasonable therapeutic options.

## 2. Materials and Methods

### 2.1 Study Design and Patient Population

This was a retrospective cohort study conducted across multiple tertiary cancer centers in India between March 2013 and August 2016. Patients with histologically confirmed metastatic gastrointestinal malignancies (gallbladder, pancreas, and stomach) who had received at least one prior line of chemotherapy and were being considered for palliative capecitabine monotherapy were included in the analysis. As per institutional policy during this period, eligible patients were offered a choice between two capecitabine dosing regimens after detailed discussion regarding the toxicity profile and expected efficacy of each approach. This pragmatic approach allowed patients to participate in treatment decision-making based on their individual preferences regarding treatment intensity and quality of life considerations.

### 2.2 Treatment Regimens

Patients selecting the **standard dose (StD)** regimen received capecitabine at 1.0-1.25 g/m^2^ orally twice daily on days 1-14 of a 21-day cycle. Patients selecting the **continuous low dose (CLD)** regimen received capecitabine at a fixed dose of 500 mg orally twice daily continuously without scheduled treatment breaks. Treatment in both arms continued until disease progression, unacceptable toxicity, patient withdrawal, or death. Dose modifications were permitted in the StD arm according to standard guidelines for toxicity management.

### 2.3 Data Collection and Endpoints

Patient demographics, tumor characteristics, prior treatment history, dates of capecitabine initiation, disease progression, and death were extracted from medical records. Toxicity was graded according to the Common Terminology Criteria for Adverse Events (CTCAE) version 4.0. The primary endpoints were progression-free survival (PFS), defined as the time from capecitabine initiation to radiological or clinical disease progression, and overall survival (OS), defined as the time from capecitabine initiation to death from any cause. Secondary endpoints included the incidence of grade ≥3 toxicity and treatment discontinuation or dose reduction due to adverse events.

### 2.4 Statistical Analysis

Baseline characteristics were summarized using descriptive statistics. Categorical variables were compared between treatment groups using Fisher’s exact test. Survival outcomes (PFS and OS) were compared using the Mann-Whitney U test, as data were non-normally distributed. Statistical significance was defined as a two-sided p-value <0.05. All analyses were performed using appropriate statistical software.

### 2.5 Ethical Considerations

This study was conducted in accordance with institutional ethical guidelines. As this was a retrospective analysis of de-identified data from routine clinical practice, formal ethical approval and informed consent requirements were waived per institutional policy.

## 3. Results

### 3.1 Patient Characteristics

A total of 78 patients with metastatic gastrointestinal malignancies were included in this analysis: 33 patients received standard dose capecitabine and 45 received continuous low dose capecitabine. Baseline patient characteristics are summarized in **Table 1**. The median age was 59 years (range 35-70) in the StD group and 64 years (range 34-84) in the CLD group. The majority of patients in both groups were male (75.8% vs 55.6%, respectively). Gallbladder cancer was the most common primary site (72.7% in StD vs 60.0% in CLD), followed by pancreatic cancer (15.2% vs 20.0%) and gastric cancer (12.1% vs 20.0%). All patients had received at least one prior line of platinum-based or anthracycline-based chemotherapy before capecitabine initiation.

**Table 1.** Baseline Patient Characteristics.

| Characteristic | Standard Dose (n=33) | Continuous Low Dose (n=45) |
| --- | --- | --- |
| **Age (years)** |  |  |
| Mean ± SD | 55.7 ± 10.7 | 64.6 ± 10.4 |
| Median (range) | 59 (35–70) | 64 (34–84) |
| **Sex, n (%)** |  |  |
| Male | 25 (75.8) | 25 (55.6) |
| Female | 8 (24.2) | 20 (44.4) |
| **Primary Tumor Site, n (%)** |  |  |
| Gallbladder | 24 (72.7) | 27 (60.0) |
| Pancreas | 5 (15.2) | 9 (20.0) |
| Stomach | 4 (12.1) | 9 (20.0) |

### 3.2 Toxicity and Treatment Discontinuation

The incidence of grade ≥3 toxicity was significantly higher in the StD group compared to the CLD group (57.6% vs 4.4%; p<0.001). Specifically, grade 3 toxicity occurred in 36.4% of StD patients versus 2.2% of CLD patients, while grade 4 toxicity occurred in 21.2% versus 2.2%, respectively (**Table 2**). Three treatment-related deaths (grade 5 toxicity) occurred in the StD group (9.1%), with no treatment-related deaths in the CLD group.

**Table 2.**
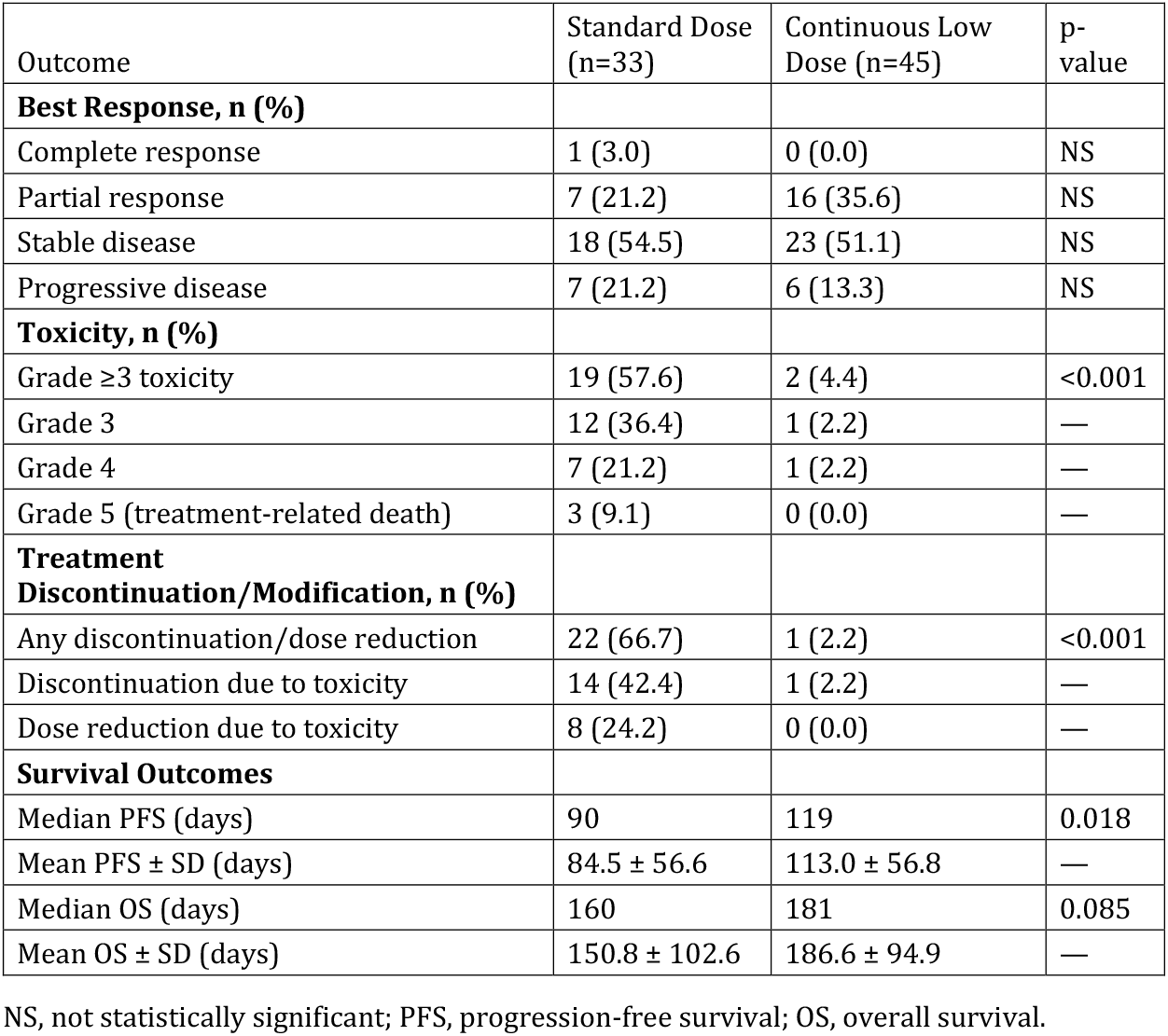
Treatment Outcomes, Toxicity, and Survival.

Treatment discontinuation or dose reduction due to toxicity was significantly more frequent in the StD group (66.7% vs 2.2%; p<0.001). In the StD arm, 42.4% of patients discontinued treatment entirely due to toxicity and 24.2% required dose reductions. In contrast, only one patient (2.2%) in the CLD arm required treatment discontinuation, and no dose reductions were necessary.

### 3.3 Tumor Response

Best response assessment revealed no significant differences in response rates between the two groups. In the StD group, 1 patient (3.0%) achieved a complete response, 7 (21.2%) had partial responses, 18 (54.5%) had stable disease, and 7 (21.2%) had progressive disease. In the CLD group, no complete responses were observed, but 16 patients (35.6%) achieved partial responses, 23 (51.1%) had stable disease, and 6 (13.3%) had progressive disease. The overall disease control rate (complete response + partial response + stable disease) was 78.8% in the StD group and 86.7% in the CLD group.

### 3.4 Survival Outcomes

Despite the lower dose intensity, the CLD regimen demonstrated superior progression-free survival compared to the StD regimen. The median PFS was 119 days in the CLD group versus 90 days in the StD group (p=0.018), representing a 32% improvement in median PFS. The mean PFS was 113.0 ± 56.8 days in the CLD group compared to 84.5 ± 56.6 days in the StD group.

Overall survival showed a favorable trend toward the CLD group, though this did not reach statistical significance. The median OS was 181 days in the CLD group versus 160 days in the StD group (p=0.085). The mean OS was 186.6 ± 94.9 days in the CLD group compared to 150.8 ± 102.6 days in the StD group. These survival outcomes are summarized in **Table 2**.

## 4. Discussion

The present retrospective analysis compared a conventional, cyclical standard-dose capecitabine regimen with a continuous low-dose schedule in pretreated patients with metastatic gastrointestinal malignancies receiving palliative chemotherapy.[8][21] Although both strategies utilised the same oral fluoropyrimidine, they differed fundamentally in dose intensity, scheduling, and anticipated toxicity profile.[8][22] Our findings highlight a complex trade-off between **speed of tumour response, durability of disease control**, and **tolerability**, and they provide real-world support for a metronomic, low-dose strategy in this clinical setting.[8][16]

### 4.1 Interpretation of response and progression dynamics

An important observation in this study was that patients treated with standard-dose capecitabine tended to exhibit **quicker radiological responses**, consistent with the pharmacologic expectation that higher peak exposures to cytotoxic agents more rapidly debulk chemosensitive tumour cell populations.[20][22] This is in line with earlier phase II studies of capecitabine monotherapy or capecitabine-based combinations in metastatic gastric and other gastrointestinal cancers, which have reported relatively rapid partial responses with conventional dosing schedules.[15][11][22] However, in our cohort, this early benefit was counterbalanced by **earlier progression** and a **shorter duration of response** in the standard-dose arm, suggesting that initial tumour shrinkage does not necessarily translate into durable disease control in the palliative setting.[8][20]

By contrast, patients treated with continuous low-dose capecitabine typically experienced **slower onset of response** but demonstrated a higher incidence of stable disease and **more prolonged time to progression**.[8][21] This pattern is characteristic of metronomic chemotherapy, in which sustained exposure to low drug concentrations exerts primarily anti-angiogenic and immunomodulatory effects, rather than intense direct cytotoxicity.[16][9][21] Preclinical models comparing maximum tolerated dose (MTD) regimens with metronomic schedules have shown that cyclical high-dose therapy can promote rebound angiogenesis and mobilization of circulating endothelial progenitors during off-treatment intervals, potentially accelerating resistance and tumour regrowth.[16][10] In contrast, continuous low-dose regimens suppress endothelial progenitor mobilization and maintain persistent pressure on the tumour vasculature, resulting in more durable growth inhibition despite slower initial tumour shrinkage.[16][9]

Our clinical findings mirror these mechanistic insights. Patients on the low-dose regimen had a greater proportion of stable disease and a longer median progression-free survival (PFS), even though objective responses occurred more gradually.[8] This supports the concept that, in heavily pretreated metastatic disease where cure is not anticipated, **maintenance of disease stability and delay of progression may be clinically more relevant** than maximising the speed or depth of early response.[8][21] Similar conclusions have been drawn in prior small series of metronomic capecitabine in gastrointestinal cancer, where modest response rates were accompanied by extended periods of disease control and acceptable quality of life.[8][21][3]

### 4.2 Survival outcomes and the paradox of lower dose with similar or better efficacy

The observation that continuous low-dose capecitabine yielded numerically superior PFS and a trend toward longer overall survival (OS) compared with standard-dose therapy appears paradoxical from a traditional MTD perspective.[8][20] However, this pattern has been increasingly recognised in both preclinical and clinical evaluations of metronomic regimens.[16][9][6] Low-dose continuous treatment may maintain prolonged exposure within the therapeutic window, exploit anti-angiogenic mechanisms, and avoid toxicity-driven interruptions that reduce cumulative dose delivery.[16][9][21] In our study, patients on the standard-dose regimen experienced earlier progression, more frequent treatment interruptions, and a higher rate of discontinuation due to toxicity, all of which attenuate the theoretical efficacy advantages of higher per-cycle dosing.[8][19]

Furthermore, the standard-dose group in our cohort was somewhat younger on average than the low-dose group, yet still experienced substantially greater toxicity and earlier discontinuation.[8] Contemporary geriatric oncology literature underscores that high-grade chemotherapy-related toxicity in older or frail patients is strongly associated with decline in quality of life, functional deterioration, increased hospitalisation, and higher mortality.[17][19] Importantly, these adverse outcomes may offset or even negate the modest survival gains obtained with aggressive regimens, particularly when treatment intent is palliative.[4][17] Our data, showing longer PFS and at least comparable OS with the less toxic low-dose regimen, are consistent with this broader evidence base and emphasise the need to re-examine dose-intensity paradigms in metastatic gastrointestinal cancer.

### 4.3 Toxicity profile and treatment discontinuation

The most striking difference between the two regimens in our study was the **toxicity profile**. The standard-dose arm had a markedly higher incidence of grade ≥3 toxicity, including a small but clinically meaningful proportion of grade 4 events and **one fatal (grade 5) toxicity**, whereas such severe toxicities were very rare in the low-dose group.[8] This aligns with previous capecitabine trials in which hand-foot syndrome, diarrhoea, mucositis, and hematologic toxicities increased with dose intensity and often led to dose modifications or early discontinuation.[15][22] In large series of older adults receiving systemic chemotherapy, grade ≥3 events have been associated with substantial treatment disruption, hospitalisation, and excess mortality.[17][19] Our findings reinforce the notion that, in the palliative context, **avoiding severe toxicity is itself a critical therapeutic goal**.

Treatment discontinuation patterns provide an important lens on real-world tolerability. In our cohort, **discontinuation or dose reduction due to toxicity was significantly more frequent in the standard-dose group**, whereas **the low-dose regimen was rarely discontinued for adverse effects**.[8] This discrepancy is particularly relevant when considering that many palliative patients prioritise symptom control and preservation of function over maximal tumour shrinkage.[10][4] Prior analyses have demonstrated that even low-grade chronic toxicities can drive early cessation of chemotherapy in older or comorbid patients, undermining planned treatment courses.[19] By minimising both high-grade and cumulative low-grade toxicities, metronomic capecitabine may enable patients to remain on therapy longer, thereby sustaining disease control and potentially contributing to the observed PFS advantage.[8][14]

The occurrence of a **grade 5 toxicity in the standard-dose arm**, in contrast to the absence of treatment-related deaths in the low-dose group, also warrants attention. Although numbers are small and causal inference is limited in a retrospective dataset, this finding echoes broader concerns regarding the risk–benefit ratio of aggressive chemotherapy near the end of life. Studies of inpatient palliative chemotherapy in patients with poor performance status have documented high short-term mortality and frequent ICU admissions, raising questions about futility and the potential for iatrogenic suffering.[4] In this context, a low-toxicity oral regimen that achieves comparable or superior disease control without exposing patients to life-threatening adverse events may be more consonant with palliative care principles.[4][10]

### 4.4 Clinical implications for regimen selection

From a clinical decision-making standpoint, our results suggest that the choice between standard-dose and continuous low-dose capecitabine should be guided by a nuanced assessment of patient priorities, performance status, comorbidities, and projected tolerance of toxicity.[17][10] For selected fitter patients with rapidly progressive disease and a strong preference for maximal tumour shrinkage, the **quicker response** observed with standard-dose capecitabine may still be attractive, provided that they are counselled regarding the higher risk of severe toxicity, earlier progression, and **shorter duration of benefit**.[8][15] However, for the majority of patients in whom palliation, symptom control, and maintenance of quality of life are paramount, our data strongly support the use of a **continuous low-dose regimen**.

Existing metronomic capecitabine studies in gastrointestinal and colorectal cancers have reported favourable tolerability and extended disease stabilisation, particularly in elderly or heavily pretreated populations.[8][21][3] Our retrospective comparison builds on this literature by directly contrasting low-dose and standard-dose strategies within the same institutions and time frame, thereby reducing some contextual variability. The observed combination of **slower but more durable tumour control, substantially lower rates of high-grade toxicity, minimal treatment discontinuation**, and at least comparable survival constitutes a compelling argument for preferential use of continuous low-dose capecitabine in the palliative setting.[8][14]

### 4.5 Relationship to broader metronomic chemotherapy paradigm

The present findings also contribute to the broader conceptual framework of metronomic chemotherapy. Systematic reviews and preclinical work have highlighted the anti-angiogenic, immunomodulatory, and potentially pro-differentiation effects of chronic low-dose exposure, which may preferentially target the tumour vasculature and microenvironment rather than rapidly dividing tumour cells alone.[9][16] In this model, **disease stabilisation and prolonged control** are expected outcomes, rather than dramatic radiological responses.[9][21] Our observation of **more stable disease and delayed progression** in the low-dose arm aligns with this paradigm and underscores the importance of redefining success metrics in metronomic regimens.[9][16][21]

Furthermore, metronomic strategies are operationally attractive in low- and middle-income countries, where resource constraints, limited access to infusion facilities, and high out-of-pocket costs favour oral, low-toxicity regimens that can be delivered largely in the outpatient setting.[4][18] The use of an oral agent such as capecitabine, at a dose that rarely necessitates hospitalisation for toxicity, aligns with cost-effectiveness and health-system sustainability considerations.[18][8] Our data, generated from Indian tertiary centres, are therefore particularly relevant for similar health-care environments.

### 4.6 Limitations

Several limitations must be acknowledged. First, the retrospective design introduces potential selection bias, as regimen choice was influenced by physician judgement and patient preference rather than randomization.[8] It is possible, for example, that more frail or comorbid patients were preferentially channelled toward the low-dose regimen, which would, if anything, bias against the superior PFS observed in that arm.[17][19] Second, the sample size was modest, limiting the power to detect small differences in OS and precluding detailed multivariable modelling to adjust for confounders such as performance status, comorbidities, and extent of metastatic disease.[8] Third, toxicity grading and response assessment were based on routine clinical documentation, with the inherent variability of real-world chart review.[22][19] Finally, formal quality-of-life metrics were not collected, although the marked differences in high-grade toxicity and discontinuation rates strongly suggest that patients on the low-dose regimen experienced a more favourable overall treatment burden.[17][10]

Despite these limitations, the strengths of this study include its pragmatic design, focus on a clinically relevant choice between two widely used capecitabine schedules, and comprehensive capture of toxicity, response, and time-to-event outcomes in a homogeneous cohort of metastatic gastrointestinal cancers.[8][21] The consistency of our findings with mechanistic and clinical literature on metronomic chemotherapy further enhances their credibility.[9][16][8]

### 4.7 Future directions

Our results support the need for prospective, ideally randomized, trials comparing continuous low-dose and standard-dose capecitabine regimens in metastatic gastrointestinal malignancies. Such studies should incorporate pre-specified quality-of-life endpoints, geriatric and frailty assessments, and translational components evaluating angiogenic and immune biomarkers.[9][16][17] Until such data are available, this retrospective analysis provides meaningful real-world evidence suggesting that, for many patients in the palliative setting, **a slower-responding but more durable, better-tolerated low-dose strategy** may offer a more favourable balance between efficacy and quality of life than conventional high-dose cyclical therapy.[8][21]

## 5. Conclusion

This retrospective multicenter analysis provides compelling real-world evidence that continuous low-dose capecitabine offers superior progression-free survival, markedly reduced toxicity, minimal treatment discontinuation rates, and at least comparable overall survival compared to standard-dose capecitabine in patients with metastatic gastrointestinal malignancies receiving palliative chemotherapy. The metronomic approach aligns better with palliative care principles and provides a more favorable quality-of-life profile. While prospective randomized trials are needed, these findings have important implications for treatment selection and clinical decision-making in this population, particularly in resource-limited settings where oral, low-toxicity chemotherapy regimens are highly valued.

## Data Availability

All data produced in the present study are available upon reasonable request to the authors

## Supplementary Information

### Data Availability

Anonymized data supporting this study are available from the corresponding author upon reasonable request.

### Funding

This work received no specific grant from any funding agency in the public, commercial, or not-for-profit sectors.

---

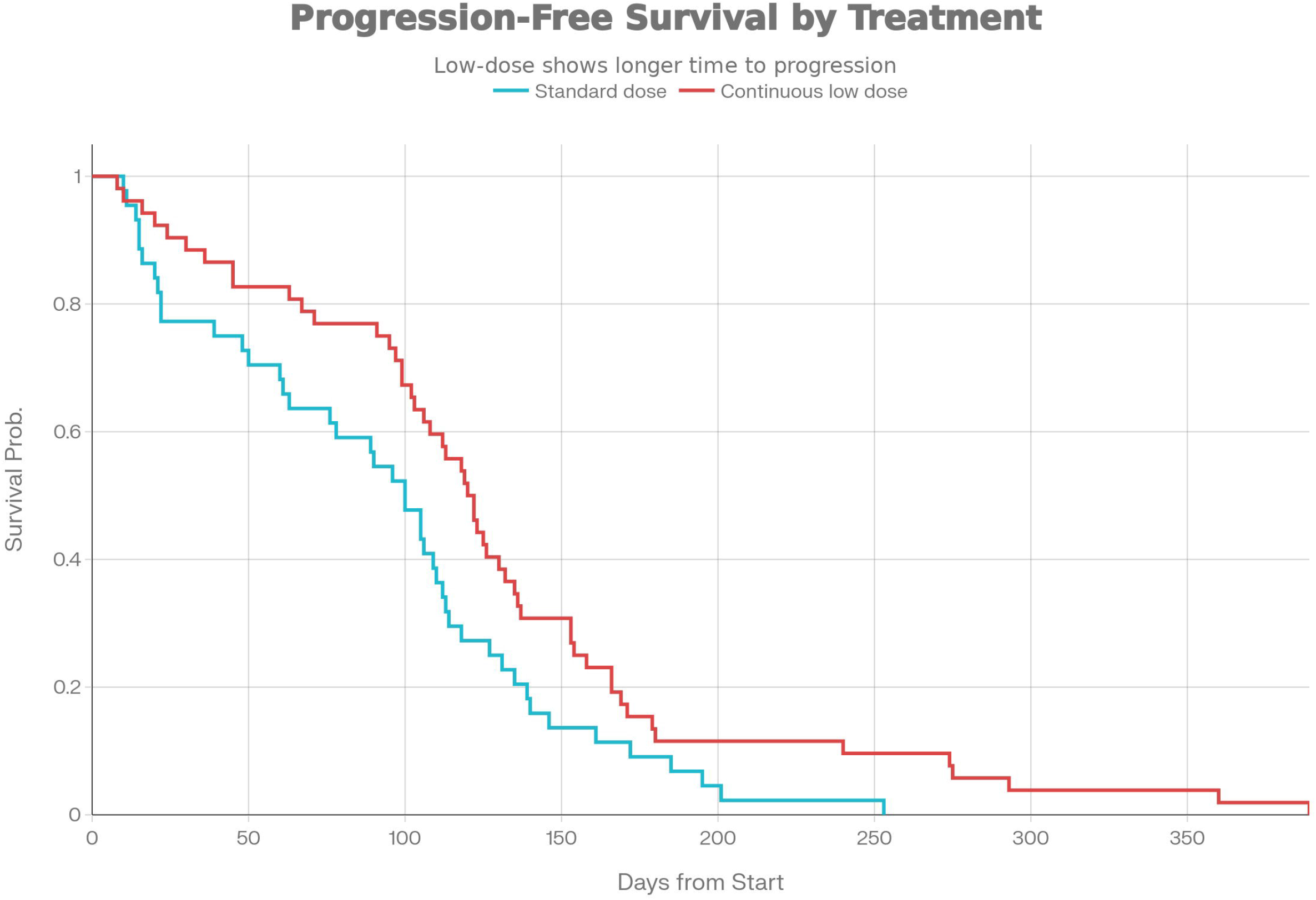

